# Science Map of the Highly Tweeted Endodontic Articles

**DOI:** 10.1101/2020.03.29.20046755

**Authors:** Jafar Kolahi, Saber Khazaei, Pedram Iranmanesh

**Affiliations:** Independent Research Scientist, Associate Editor of Dental Hypotheses, Isfahan, Iran; Department of Endodontics, School of Dentistry, Kermanshah University of Medical Sciences, Kermanshah, Iran; Department of Endodontics, Dental Research Center, Dental Research Institute, School of Dentistry, Isfahan University of Medical Sciences, Isfahan, Iran

**Keywords:** Bibliometric, endodontics, publication, social media

## Abstract

**Introduction:** To identify active journals, authors, institutions and hot topics in the field of Endodontics within the Twittersphere.

**Methods:** On December 23, 2019, the Altmetric database was searched using the titles of 11 endodontic journals. The bibliometric data of the top 5% of endodontic articles with the highest tweets number were extracted from the Web of Science and analyzed.

**Results:** Overall, 3,918 tweets (from 3,881 individual posts) related to endodontic articles from seven journals were identified, which were mostly from the U.S. The Journal of Endodontics received the most tweets. Systematic review and apical periodontitis were the most popular keywords. At the author level, Dummer PMH and Patel S and at institution level King’s College London and Cardiff University, had the largest number of popular articles in the Twittersphere. The number of tweets was not correlated with citations (r=0.007, P=0.929). No statistically significant differences were found between open access (n=41, Mean=11.19) and non-open access (n=136, Mean=9.38) articles regarding the number of tweets (P=0.648).

**Conclusions:** In the ‘Twittersphere’, the overall activity of endodontic journals and associations were low. They could be more active and increase their visibility and social impact by immediately sharing research outcomes and communicating with peers, practitioners, and patients.

## Introduction

Altmetrics is growing rapidly and acts as a complement to traditional citation-bases metrics (1,2). One of the most important altmetric resources is Twitter, which provides real-time data on mentions. Twitter is a microblogging and social networking service based in San Francisco, California, USA that was created in 2006. As of January 2020, Twitter had 340 million active users from all over the world (3) and the platform thus plays a significant role in the discovery of scholarly information and cross-disciplinary dissemination of knowledge (4). To demonstrate the power of Twitter in terms of the dissemination of scientific articles ‘The spread of true and false news online” would be a good example (5). This article was associated with 9099 tweets from 8323 users, with an upper bound of 39,938,946 followers.

Twitter is used widely by the scientific community and as a result new terms have been developed such as Twitter science stars (6) and the Kardashian index (which measures over-activity of researchers in Twittersphere) (7). This social media is used successfully by clinicians to communicate with peers and provide trusted health information to educate patients (8,9). A good example would be @MayoClinic with 49.3K tweets and 1.9M followers.

Twitter was also successfully used in scientific conferences as a peer-to-peer platform for scientific communication to present findings, exchange ideas, and develop networks. Twitter conferences could reduce the large financial costs for contributors and high environmental costs in the form of carbon emissions (10).

The number of tweets can predict highly-cited articles within the first 3 days of their publication (11). Journals with their own Twitter account get 34 percent more citations and 46 percent more tweets than journals without a Twitter account (12). Moreover, within general and internal medicine journals, the number of Twitter followers is significantly correlated with citations and impact factor (13).

Recent surveys have revealed that Twitter was the most popular social media and altmetric resource among the dental research community (14,15). In addition, Twitter is the most popular social media platform in Endodontology (16). Hence, we aimed to analyze highly tweeted endodontic articles to identify active journals, authors and hot topics. Furthermore, we planned to examine the correlation and association between the number of tweets and various bibliometric factors such as citations.

## Methods

The Altmetric database (Altmetric LLP, London, UK) was searched on 23 December, 2019 using the titles of 11 endodontic journals including Australian Endodontic Journal (ISSN: 1329-1947), Endodoncia (ISSN: 0071-0261), Endodontic Practice (ISSN:1465-9417), Endodontic Practice Today (ISSN:1753-2809), Endodontic Topics (ISSN: 1601-1538), Endodontology (ISSN:0970-7212), Evidence-Based Endodontics (ISSN: 2364-9526), International Endodontic Journal (ISSN: 0143-2885), Iranian Endodontic Journal (ISSN: 1735-7497), Journal of Endodontics (ISSN: 0099-2399) and Saudi Endodontic Journal (ISSN: 1658-5984). Bibliometric data of the top 5% endodontic articles with the highest number of tweets were extracted from the Web of Science and analyzed and visualized by means of author keyword co-occurrence, co-authorship and co-citation network analysis using VOSviewer 1.6.13 software (http://www.vosviewer.com/, Leiden University Centre for Science and Technology Studies).

The Pearson coefficient was used to analyze correlation between number of tweets, Altmetric score, Mendeley readers and citations. The Student T test was used to assess differences between open access and non-open access articles regarding number of tweets. Data analysis was carried out using R 3.6.3 software (R Foundation for Statistical Computing, Vienna, Austria). Excel 2016 was used to draw graphs and data-bar visualizations.

## Results

In total, 3,918 tweets (from 3,881 individual posts) related to endodontic articles from seven journals were located (Fig. 1), mostly from the US (Table 1). ‘Acetaminophen Old Drug, New Issues” with 279 tweets (from 47 users, with an upper bound of 186,968 followers) was the most popular endodontic article in the Twittersphere (Table 2). @endogarrido (Roman Martinez, 1113 followers and 4676 tweets) with 560 mentions related to endodontic articles was the most active Twitter account, followed by @autismepi (Ann Bauer, 2480 followers and 36100 tweets) with 218 mentions and @Dddent2 (dddent.com, 8994 followers and 62800 tweets) with 159 mentions.

**Figure 1.**
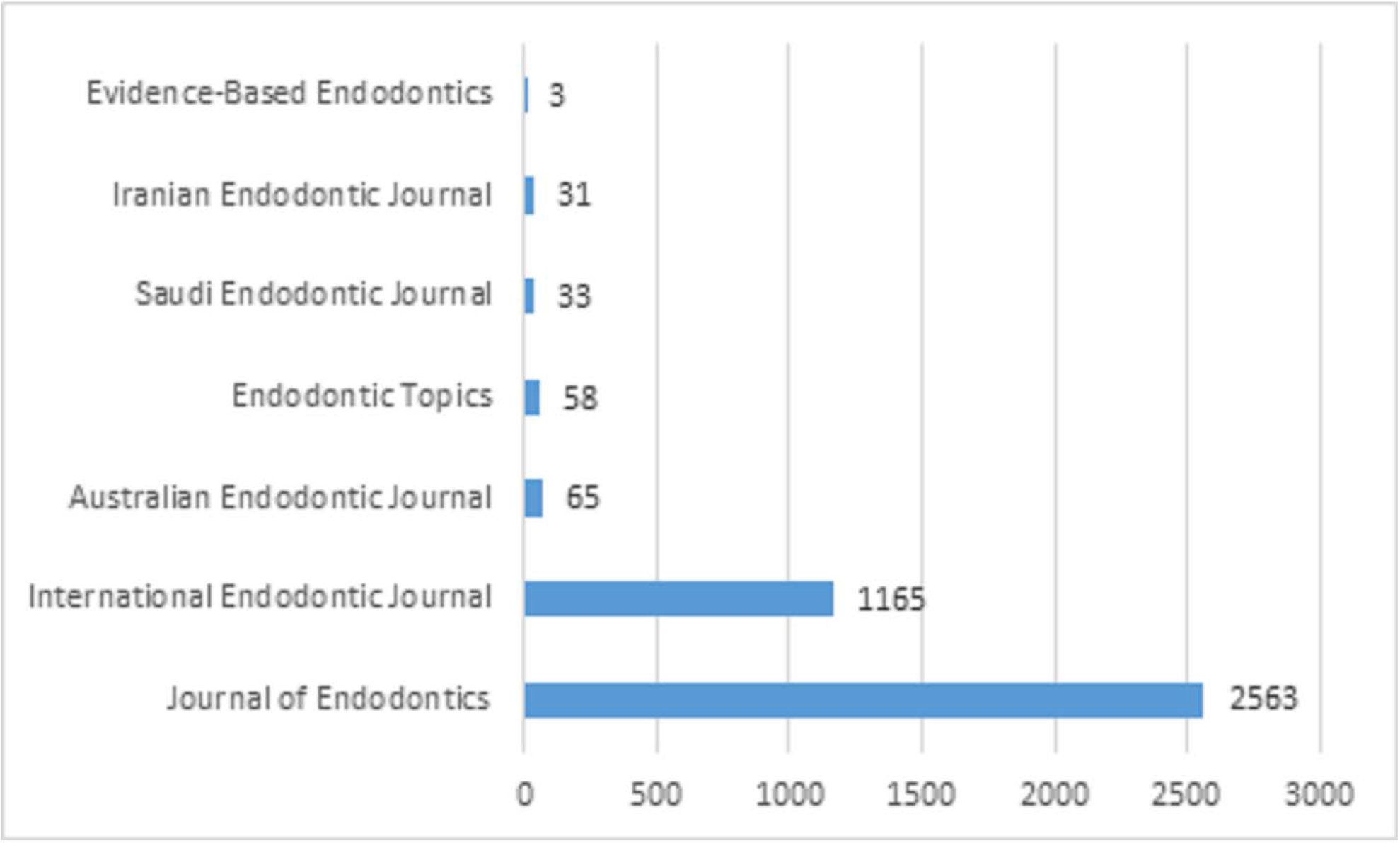
Number of Tweets among endodontic journals

**Table 1.** Top 10 countries with the highest number of Tweets related to endodontic articles

| Country | Total number of tweets | Unique tweets |
| --- | --- | --- |
| United States | 939(24%) | 151(13.5%) |
| Mexico | 575(14.7%) | 11(1%) |
| Saudi Arabia | 375(9.6%) | 181(16.2%) |
| United Kingdom | 140(3.6%) | 53(4.8%) |
| India | 112(2.9%) | 10(0.9%) |
| Canada | 94(2.4%) | 18(1.6%) |
| Norway | 94(2.4%) | 1(0.1%) |
| Spain | 68(1.7%) | 40(3.6%) |
| Chile | 63(1.6%) | 10(0.9%) |
| Greece | 41(1%) | 3(0.3%) |

**Table 2.** Top 10 endodontic articles with the highest number of Tweets

| **Number of tweeds** | **Title of article** | **Title of journal** | **Year of publication** | **Score of Altmetric** | **Number of citations** |
| --- | --- | --- | --- | --- | --- |
| 279 | Acetaminophen: old drug, new issues | Journal of Endodontics | 2015 | 30 | 31 |
| 67 | European Society of Endodontology position statement: the use of antibiotics in endodontics | International Endodontic Journal | 2018 | 41 | 26 |
| 65 | Contamination during Endodontic Treatment Is One of the Sources of Nosocomial Endodontic Propionibacterium acnes Infections | Journal of Endodontics | 2016 | 53 | 7 |
| 59 | Comparison of Cyclic Fatigue Resistance of 5 Heat-treated Nickel-titanium Reciprocating Systems in Canals with Single and Double Curvatures | Journal of Endodontics | 2019 | 50 | 0 |
| 30 | Antibiotics in Endodontics: a review | International Endodontic Journal | 2017 | 22 | 28 |
| 28 | Outcome of Partial Pulpotomy in Cariously Exposed Posterior Permanent Teeth: A Systematic Review and Meta-analysis | Journal of Endodontics | 2019 | 28 | 1 |
| 28 | European Society of Endodontology position statement: Management of deep caries and the exposed pulp | International Endodontic Journal | 2019 | 17 | 7 |
| 26 | The Effect of Long-term Dressing with Calcium Hydroxide on the Fracture Susceptibility of Teeth | Journal of Endodontics | 2018 | 22 | 4 |
| 24 | Association between Pulp Stones and Kidney Stones: A Systematic Review and Meta-analysis | Journal of Endodontics | 2019 | 18 | 1 |
| 22 | A Comparative Study of ProTaper Universal and ProTaper Next Used by Undergraduate Students to Prepare Root Canals | Journal of Endodontics | 2017 | 16 | 2 |

Source of citations: Dimensions database

Overall, the top 5% of endodontic articles with the highest number of tweets were associated with 177 articles. Journal of Endodontics had the largest number of popular articles in Twittersphere (Fig. 2).

**Figure 2.**
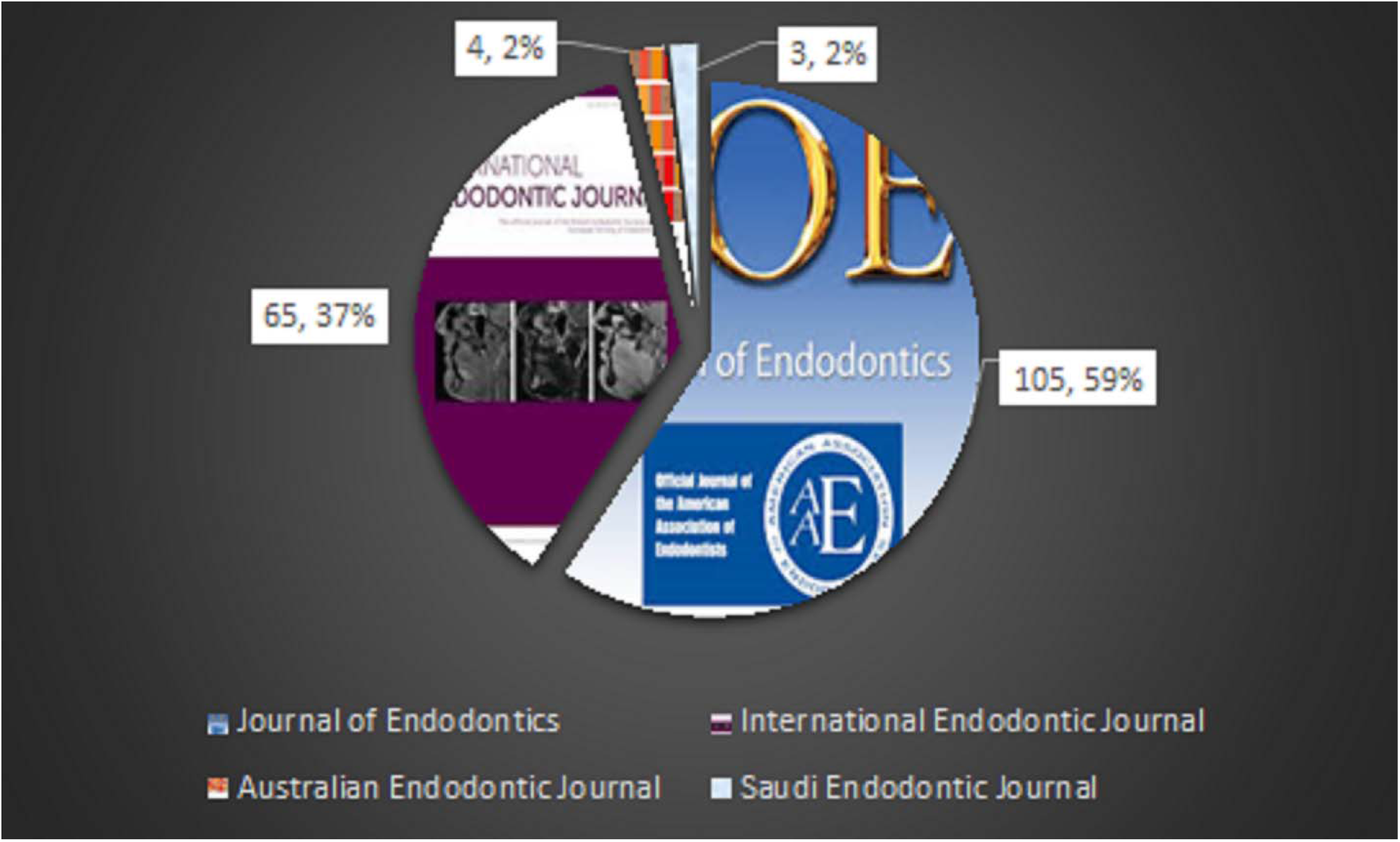
Resources of top 5% endodontic articles with the highest number of Tweets

Systematic review and apical periodontitis were the most popular keywords (Fig. 2). At author level Dummer PMH and Patel S and at institution level the King’s College London and Cardiff University, had the largest number of popular articles in the Twittersphere (Fig. 3).

**Figure 3.**
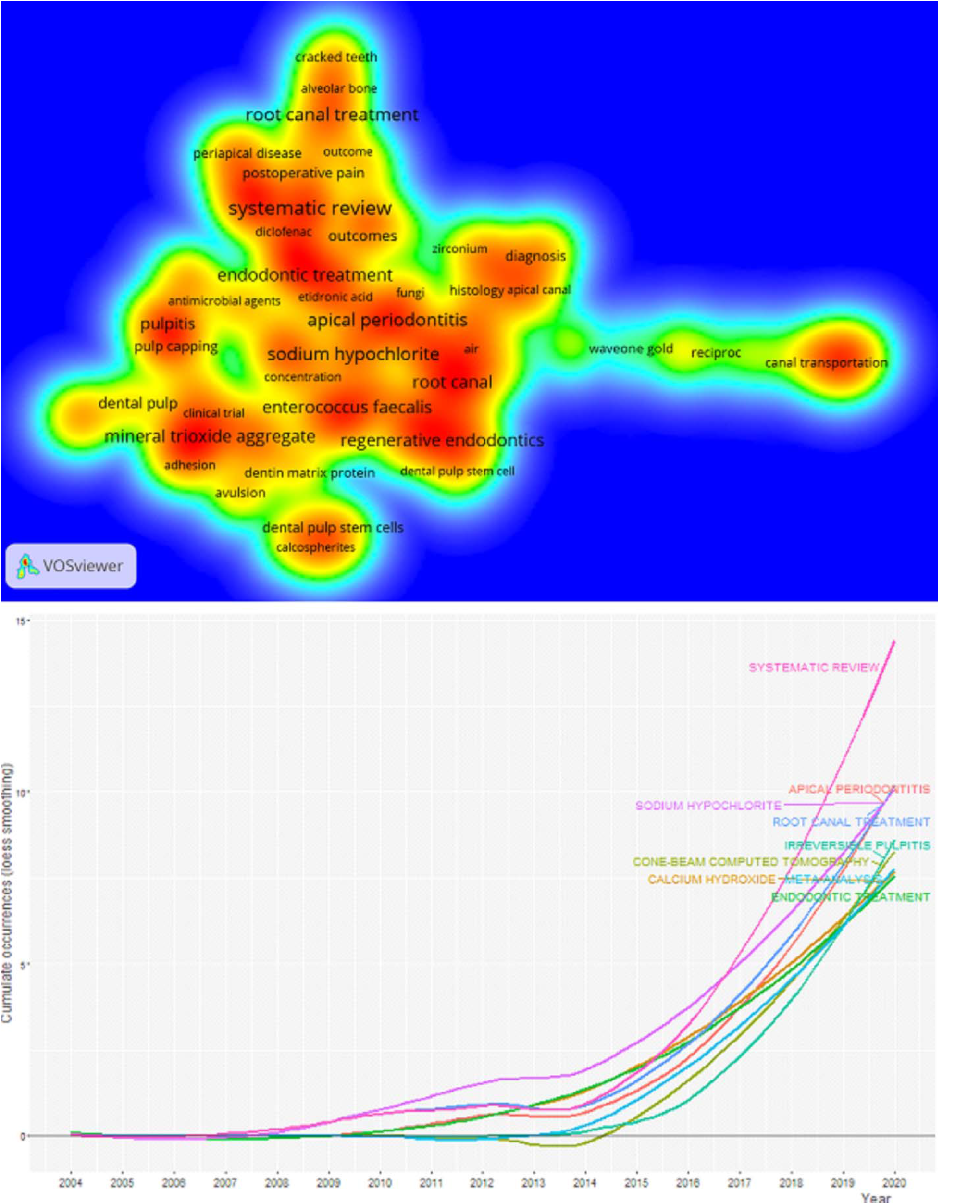
Hot topics among top 5% endodontic articles with the highest number of Tweets, created by means of author keyword co-occurrence network analysis. The lower part showed the top 10 author’s keyword growth over years.

Co-citation network analysis revealed the Journal of Endodontics had the greatest influence on the network (Fig. 4).

**Figure 4.**
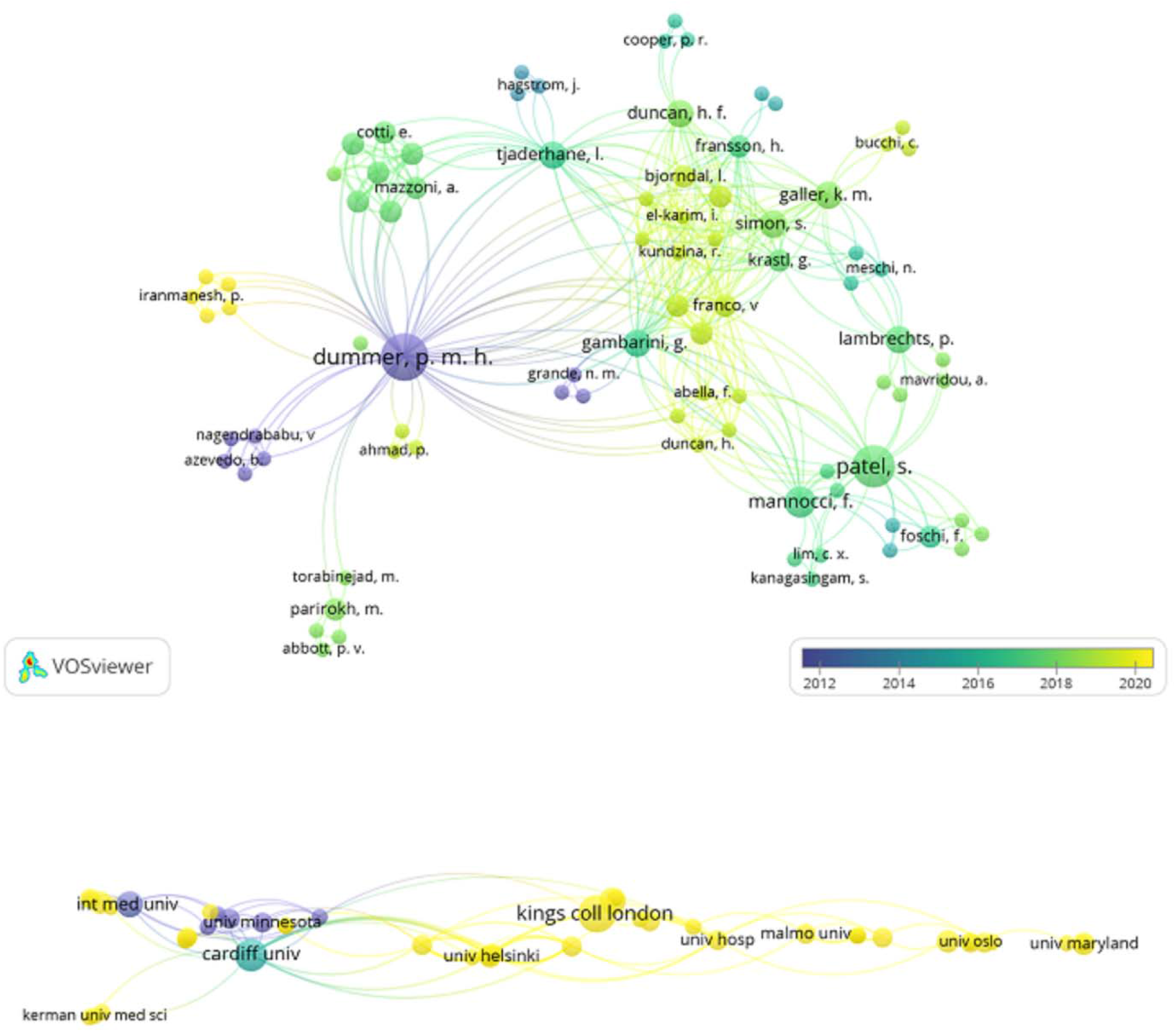
Influential authors (upper part), institutions (lower part) among top 5% endodontic articles with the highest number of Tweets, created by means of co-authorship network analysis. The color scheme showed the year of publication.

Among 177 popular articles in the Twittersphere, the number of tweets was not correlated with citations (source of citation was Dimensions database) (r=0.007, P=: 0.929), Yet, the non-parametric Locally Weighted Scatterplot Smoothing (LOWESS) line revealed for citations ranging from 20 to 40 that a relationship existed between tweets and citations (Fig. 5).

**Figure 5.**
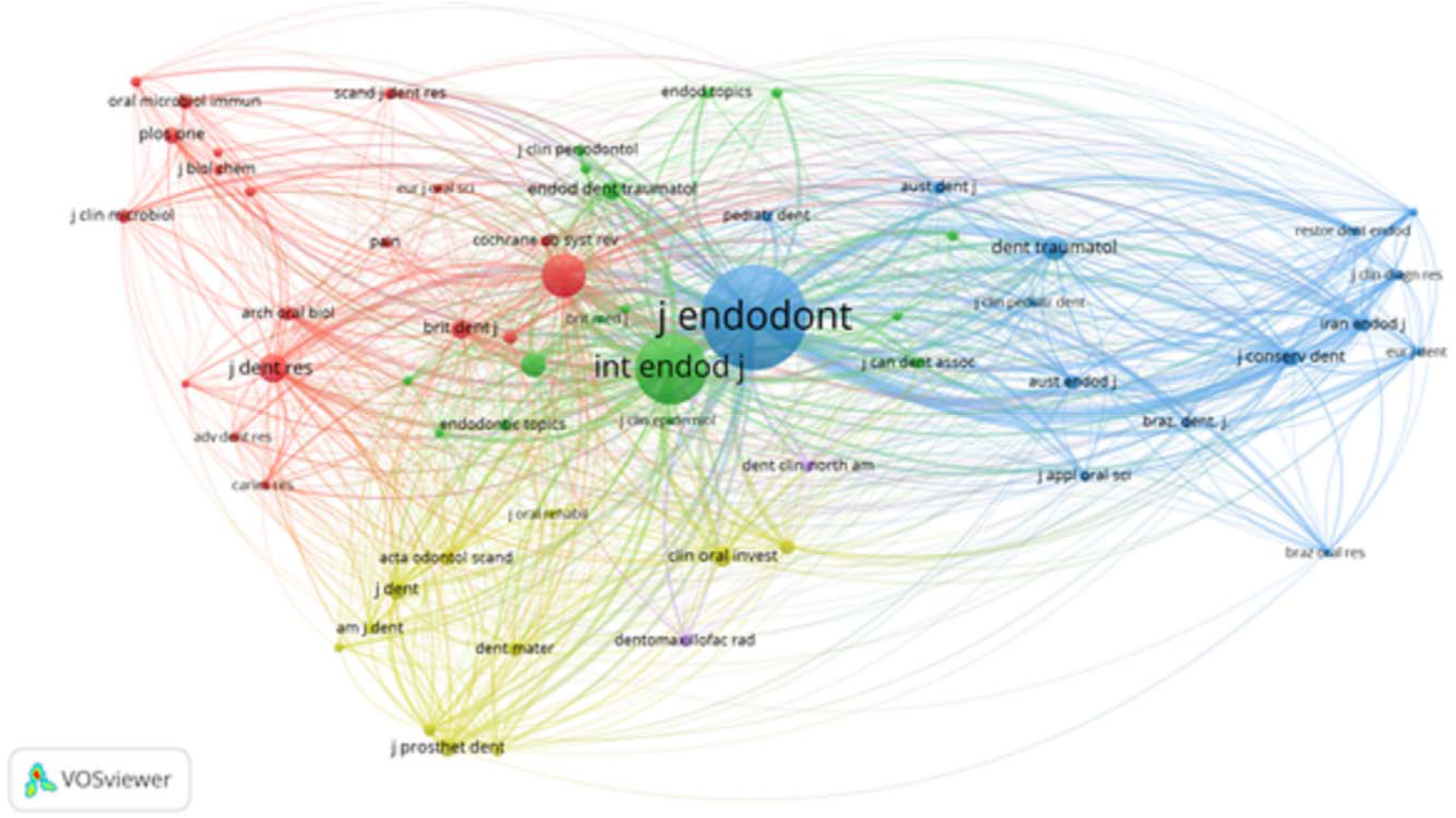
Influential resources among top 5% endodontic articles with the highest Tweets numbers, created by means of co-citation network analysis.

**Figure 6.**
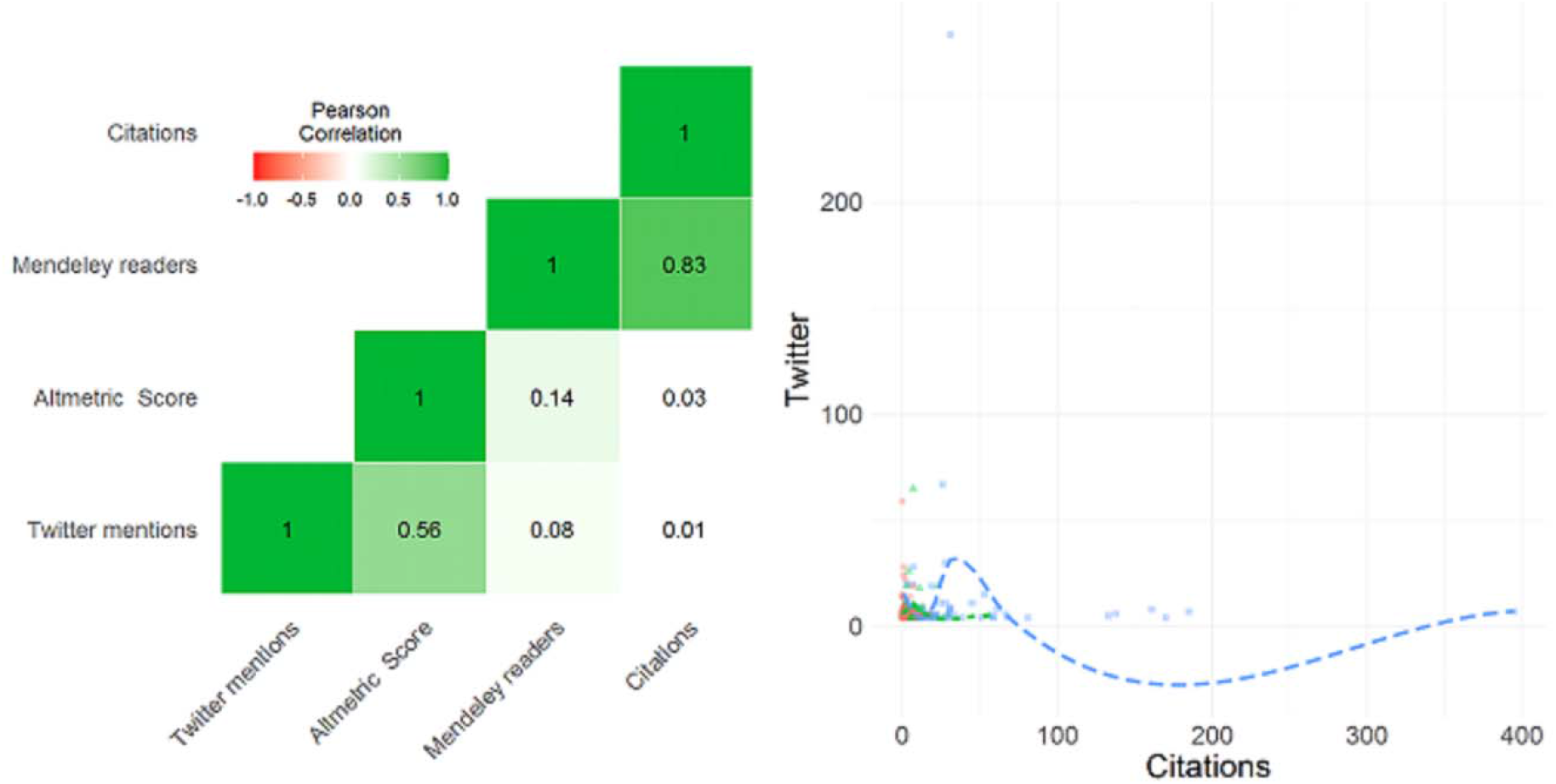
Heat Map showing the correlation between number of tweets numbers, Mendeley readers, citations and Altmetric score among top 5% endodontic articles with the highest number of tweets (n=177). Non-parametric Locally Weighted Scatterplot Smoothing (LOWESS) line examining the relationship between number of tweets and citations showed on the right side.

No statistically significant difference found between open access (n=41, Mean=11.19) and non-open access (n=136, Mean=9.38) articles regarding number of tweets (P=0.648).

## Discussion

Twitter added 5 million daily users recently (17) with a total of 2000 healthcare professionals active in the Twittersphere with at least one tweet daily (9). The endodontic community could use this free and powerful platform to create virtual communities with peers and disseminate scholarly information and research findings with members of the public.

Highly tweeted science articles and dental articles have been analyzed (18,19) and revealed that children, dental caries and periodontal diseases were hot topics among highly tweeted dental articles. The British Dental Journal, Journal of Dental Research and Journal of the American Dental Association with 39087, 4503 and 3960 tweets respectively had the largest number of tweets. The UK and US had tweeted the majority of highly tweeted dental articles. The tweeted dental articles revealed 3.9 tweetsper an article on average (19).

The Journal of Endodontics received the most tweets. This journal, the official journal of the American Association of Endodontists, was active in the Twittersphere via @savingyourteeth (4,748 tweets and 3,224 followers). The International Endodontic Journal, which is the official journal of several national endodontic societies and the European Society of Endodontology, had a lower level of activity via @EuSocEndo (474 tweets and 239 followers). This journal is ranked first among all endodontic journals according to the 2019 Journal Citation Reports (Clarivate Analytics) (Impact factor: 3.801) and could be more active in the Twittersphere. The Saudi Endodontic Journal and Saudi Endodontic Society were also active in the Twittersphere via @SaudiEndo (87 tweets and 721 followers) and @SaudiEndoS (225 tweets and 2,417 followers). Disappointingly, no other Twitter account related to endodontic journals was found. Endodontic journal editors should be aware that journals with their own Twitter account get 34 percent more citations (12). The twitter account of the British Dental Journal, @The_BDJ, with 23000 followers and 23100 tweets would be a good example.

The endeavors of an independent endodontist from Mexico, Roman Martinez, must be acknowledged; he actively tweets endodontic articles via @endogarrido (Roman Martinez, 1113 followers and 4676 tweets). Another active account was @autismepi (Ann Bauer, 2480 followers and 36100 tweets) who discussed the association between Acetaminophen and autism in children. As shown in Table 2, the most popular endodontic article in Twittersphere was about

Acetaminophen which is a popular medication in endodontics. Another account, @autismepi, tweets endodontic articles, whilst @Dddent2 (dddent.com, 8994 followers and 62800 tweets) is a commercial account for the sale of dental materials.

In terms of evaluating hot topics, systematic review was the hottest one. In view of principles of evidence-based density, this is promising and shows that in addition to editors that pay more attention to the papers with the highest level of evidence, the public view focused on these topics. This result may also be in contrast with the idea that the majority of tweets were carried out by members of the public which attracted buzzwords in the title of scientific articles. As with other reports in the field of dentistry (16,20–22), newly emerging and ground-breaking topics, e.g. genomic medicine, nanotechnology, artificial intelligence and machine learning were not included among highly tweeted endodontic articles.

In this study no statistically significant difference was found between open access and non-open access articles regarding the number of tweets. This results was in contrast with findings of a recent survey which reported Open access dental articles had significantly higher numbers of tweets (23).

The number of tweets was not correlated with citations among the top 5% endodontic articles with the highest tweets number Yet, citations were strongly correlated with the number of Mendeley readers (r= 0.8). Same lower correlation reported in dental science (r= 0.5) (24). The correlation between number of tweets and citations may be increased in future by increasing knowledge and attitude of endodontic researchers and practitioners regarding the importance of activity in the Twittersphere and other social media. As a recommendation, we suggest future evaluations will assess changes of a correlation pattern between number of tweets and citations over time.

The limitations of this study include the fact that the number of tweets may fluctuate over time. Tweets may be made from fake accounts and robots (25), indeed, it is well-known that there are spam companies who market tweets, Twitter followers and likes. In addition, censorship of Twitter in some countries with a growing number of scientific articles must be noted.

Despite the advantages of Twitter, the endodontic community must be alert about overuse of this social media phenomenon. The Kardashian index is a useful tool to measure over-activity of researchers in the Twittersphere (7). Interestingly, there were no Kardashian scientists among the endodontic community.

## Data Availability

All data is avaiable

